# Impact of commissural alignment on the hemodynamic performance of supra-annular self-expandable transcatheter aortic valves

**DOI:** 10.1101/2023.05.23.23290422

**Authors:** Ignacio J. Amat-Santos, Javier Gómez, Pablo Pinon, Luis Nombela-Franco, Raúl Moreno, Antonio J. Munoz-García, Alfredo Redondo, Antonio Gómez-Menchero, Sara Blasco-Turrión, Jorge Salgado Fernandez, Gabrsiela Tirado-Conte, Silvio Vera-Vera, Juan H. Alonso-Briales, Ramiro Trillo, Santiago Camacho Freire, Juan Pablo Sánchez-Luna, Jose Carlos Gonzalez-García, Giorgio Marengo, Christian Aristizabal, Hipólito Gutiérrez-García, Ana Serrador-Frutos, Ana Revilla, Carlos Baladrón, Manuel Carrasco-Moraleja, Itziar Gómez-Salvador, J. Alberto San Román

## Abstract

**Background:** Impact of commissural alignment (CA) strategies with self-expandable supra-annular transcatheter aortic valves (TAVR) has not been investigated yet.

**Methods:** Multicentric ambispective study including patients who underwent self-expandable TAVR in 7 centers with the Evolut Pro/Pro+ (EP)(Medtronic, USA) and Acurate neo2 (AN2)(Boston Scientific, USA). Comparison of those with CA attempt vs. those with no CA attempt was performed. The degree of commissural misalignment (CMA) was assessed by computed tomography/angiography and 1-year transvalvular gradients/regurgitation evaluated by echocardiography. A matched comparison according to annular dimensions/eccentricity, prosthesis size/type, and baseline left ventricular function and gradients was performed.

**Results:** A total of 557 patients, mean age 80.7±6.6 years, 61.4% men, and STS score of 4.3±3.1% were analyzed. A CA technique was attempted in 215 patients (38.6%), including 113 patients with AN2 and 102 patients with EP. None/mild CMA was found in 158 (73.5% vs. 43.6% if no CA attempted, p<0.001) with no differences between devices (AN2:75.2%; EP:71.6%, p=0.545). Patients with moderate/severe CMA had a greater aortic peak gradient (22.3±8.7 vs. 19.7±8.5, p=0.001), significantly greater progression of both peak (p=0.002) and mean gradients (p=0.001) after matching, and higher rate of central aortic regurgitation (1.2% vs. 0.4%, p=0.005) at 1-year, but not a greater proportion of patients with mean gradient ≥10mmHg.

**Conclusions:** The use of CA strategies significantly reduced the rate of CMA for the self-expandable TAVR devices ACN2 and EP which was associated to lower transvalvular gradients and intra-prosthetic regurgitation progression at 1-year although no criteria of structural deterioration were met at this follow up. *Clinicaltrials.org: NCT05097183*

**CONDENSED ABSTRACT:** Commissural alignment (CA) strategies with Evolut Pro and Acurate neo2 TAVR devices were similarly successful in ¾ of the patients, representing a significant increase compared to traditional implantation. Patients with moderate/severe commissural misalignment (CMA) had a significantly greater progression of both peak and mean gradients and higher rate of new central aortic regurgitation at 1-year, but not a greater proportion of patients with mean gradient above 10mmHg. Hence, even at only 1-year of follow up and despite the lack of formal structural deterioration criteria, some hints suggest a more rapid deterioration if moderate/severe CMA is present after supra-annular TAVR.

## INTRODUCTION

Despite the good results in terms of safety and efficacy of transcatheter aortic valve replacement (TAVR), its extension to younger lower risk patients has pushed the boundaries of its performance targeting for better durability and optimal results in eventual need for coronary or valvular reintervention. In particular, the alignment between the neo-commissures and the native valve commissures (commissural alignment, CA) might facilitate coronary re-access and reduce the risk of coronary obstruction in the need for TAV-in-TAV if prosthetic degeneration occurs.

Interestingly, a recent series by Raschpichler et al (1) suggests that the lack of CA in patients receiving balloon-expandable intra-annular devices is associated to a greater rate of transvalvular gradients, although prospective case series have not confirmed this end. If confirmed, a group-effect cannot be assumed, in particular when referred to supra-annular devices, whose behavior in former studies has constantly demonstrated better gradients and lower rate of prosthesis-patient mismatch than the Edwards series (2,3,4). Moreover, the use of supra-annular self-expandable Evolut R/PRO (EP) (Medtronic, USA) was an independent predictor of failed coronary catheter canulation in the RE-ACCESS study (5), suggesting particular relevance of adequate coronary alignment when this device is used.

Therefore, we aimed to explore the efficacy of CA strategies with self-expandable supra-annular devices to achieving adequate alignment in a multicenter series, and the potential impact in post-TAVR hemodynamics (including aortic regurgitation and transvalvular gradients) at long-term follow up.

## METHODS

Multicentric ambispective study including patients who underwent successful self-expandable TAVR in 7 centers in Spain with the EP and Acurate neo2 (AN2) (Boston Scientific, USA) devices who were alive at 1-year follow up. In all the institutions different CA strategies were initiated after 2019, including standardized positioning (Evolut series at 3 o’clock and AN2 at 6 o’clock) or patient-tailored orientation (ACA trial, NCT05097183). Patients treated with TAVR in 2018 and 2019 before the start of CA strategies and who had computed tomography (CT) and echocardiography available at 1-year follow up due to different research protocols were used as control group.

Degree of commissural misalignment (CMA) was evaluated after the procedure in all the included patients. The method for evaluating this included both, angiography (59.4%) and CT (40.6%) analysis as previously explained elsewhere (6) and the kappa index for the degree of agreement between both methods was estimated. According to previous consensus (7), the degree of CA was defined as the angular relationship between native and prosthesis valve commissures in four categories of misalignment: none (≤15°), mild (15.1°-30°), moderate (30.1-45.0°) and severe misalignment 45.1-60°). When CT was available, coronary alignment and the presence of any data suggesting leaflet thrombosis were also gathered. Echocardiographic parameters and clinical variables were recorded at baseline, 30-day, and 1-year follow-up. All available CT and echocardiographic images were centrally analyzed. The study was approved by local ethics committees that waived the obtention of informed consent from the retrospective cases; all patients included prospectively in the ACA trial provided informed consent and all imaging exams were centrally analyzed.

### Study endpoints

The primary endpoint was the rate of none or mild CMA in patients with CA dedicated strategies compared to those without attempt to align the prosthesis.

The secondary endpoint included the impact of the degree of CA in the hemodynamics at 1-year follow up including degree and mechanism of aortic regurgitation, and progression of transvalvular gradients.

### Statistical analysis

We reported categorical variables as absolute values and percentages. Continuous variables were reported as mean ± standard deviation (median [IQR]). T-student or U Mann Whitney tests were used for comparing continuous variables according to variable distribution (normal or not). The normal distribution of continuous variables was verified with the Kolmogorov-Smirnov test and q-q plot. Categorical variables were compared with the chi-square test and the Fisher exact test when necessary. We compared continuous variables with the Mann-Whitney U test. We performed the statistical analyses with the use of R software, version 3.6.1 (R Project for Statistical Computing). Eligible patients were matched according to: 1) aortic annulus area/diameter/perimeter/eccentricity index; 2) aortic valve calcium score; 3) left ventricular ejection fraction at baseline; 4) transaortic gradients at baseline; 5) prosthesis size/type (**Supplemental Figure 1**). Pairs of patients were derived using the greedy nearest neighbor method 1:1 with one-fifth of the SD of the logit of the propensity score as caliper. The MatchIt package (Ho, Imai, King & Stuart, 2007) was used (see **Supp.Table 1**). Differences were statistically significant when the *P*-value was < .05.

## RESULTS

### Baseline characteristics

A total of 557 patients from 7 institutions successfully treated with TAVR between 2018 and 2022 and alive at 1-year follow up were analyzed. Regarding the baseline characteristics of the patients (**Table 1**), the mean age was 80.7±6.6 years, most of them were men (n=342, 61.4%), and the surgical risk assessed by the Society of Thoracic Surgeons score was 4.3±3.1%. Mean left ventricular function was 56±12%, with peak and mean gradients at baseline of 71.1±21.8 and 43.8±14.3 mmHg, respectively. Patients were treated with two self-expandable devices: AN2 (Boston Scientic, USA) (N=301) and EP (Medtronic, USA) (N=256) with peak and mean gradients post-implantation of 20.7±8.6 and 9.4±4.3 mmHg. Main procedural and in-hospital outcomes according to the device used were similar and are summarized in **Table 2**. Procedural success was 98.4% and there were no other clinical differences in the outcomes according to the device used.

**Table 1.**
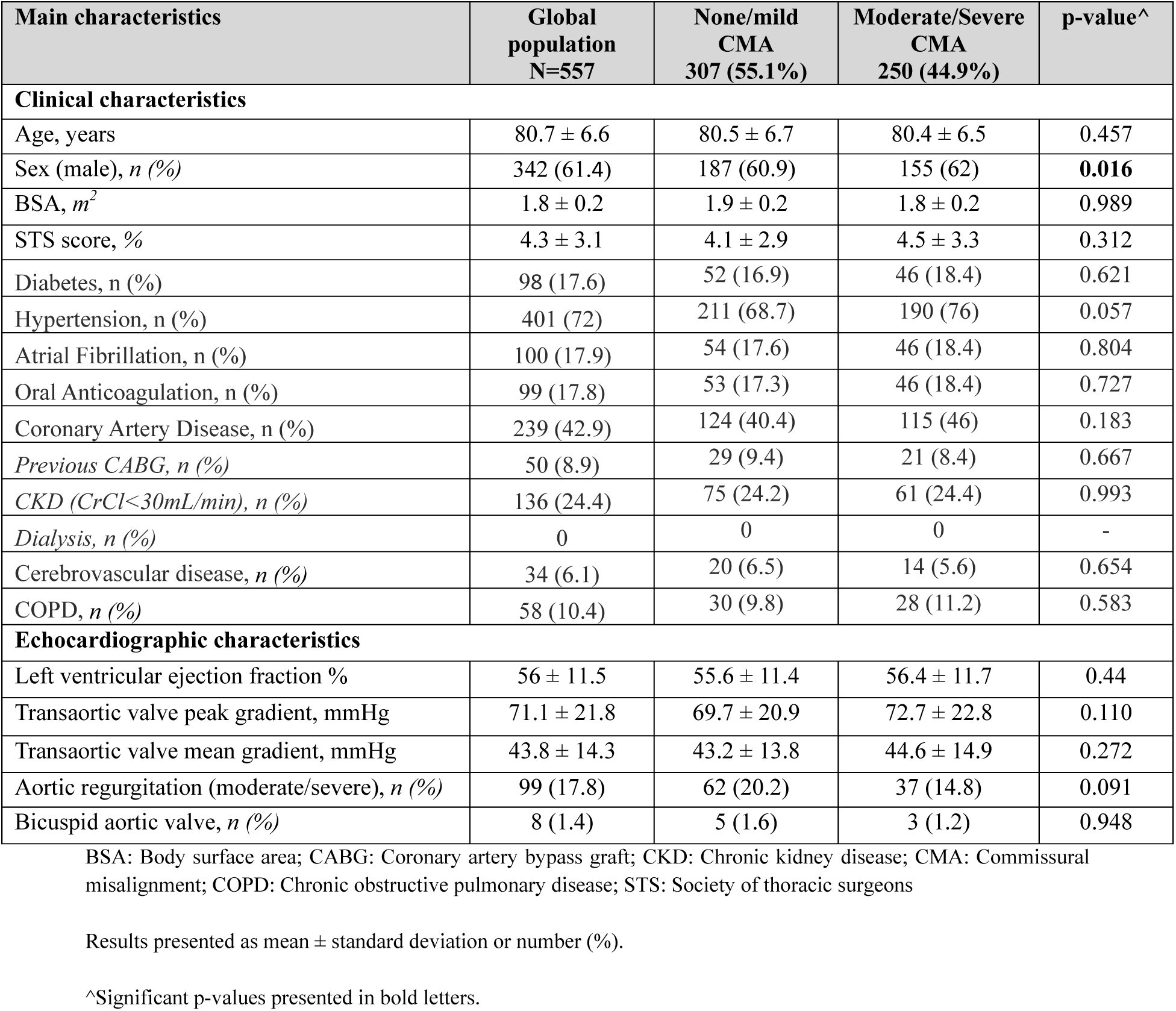
Baseline characteristics in the global study population and according to the degree of commissural misalignment.

**Table 2.** Baseline characteristics and hemodynamic performance according to the device used.

| MAIN CHARACTERISTICS | Evolut<br>N=256 | Acurate<br>N=301 | p-value<sup>^</sup> |
| --- | --- | --- | --- |
| **Baseline characteristics** |  |  |  |
| Age, years | 80.7 ± 6.8 | 80.6 ± 6.3 | 0.602 |
| BSA, m<sup>2</sup> | 1.8 ± 0.2 | 1.8 ± 0.2 | 0.999 |
| STS, % | 4.4 ± 3.10 | 4.3 ± 3.2 | 0.560 |
| Gender, n (%) male | 149 (58.2) | 193 (64.1) | 0.153 |
| Baseline left ventricular ejection fraction, % | 55 ± 10 | 56 ± 12 | 0.137 |
| Baseline peak gradient, mmHg | 70.9 ± 21.8 | 71.2 ± 21.9 | 0.901 |
| Baseline mean gradient, mmHg | 43.9 ± 14.5 | 43.7 ± 14.2 | 0.860 |
| Aortic annulus (mean diameter), mm | 25.1 ± 2.8 | 24.8 ± 2.0 | 0.259 |
| Eccentricity index, ratio | 0.18 ± 0.07 | 0.19 ± 0.08 | 0.204 |
| Aortic annulus perimeter, mm | 78.9 ± 7.8 | 75.2 ± 4.2 | **< 0.001** |
| Baseline aortic regurgitation (moderate/severe), n (%) | 52 (20.3) | 47 (15.6) | 0.148 |
| **1 year follow-up** |  |  |  |
| Global aortic regurgitation (moderate/severe), n, (%) | 23 (9) | 34 (11.3) | 0.370 |
| Central aortic regurgitation (moderate/severe), n (%) | 1 (0.4) | 2 (0.7) | 0.999 |
| Periprosthetic regurgitation (moderate/severe), n (%) | 23 (9) | 33 (11) | 0.439 |
| Transaortic valve peak gradient, mmHg | 20.1 ± 8.1 | 21.5 ± 9.1 | 0.055 |
| Transaortic valve mean gradient, mmHg | 10.6 ± 4.7 | 10.2 ± 4.5 | 0.350 |
| *Δ peak gradient, mmHg* | 0.19 ± 1.14 | 0.21 ± 1.52 | 0.875 |
| *Δ mean gradient, mmHg* | 1.21 ± 1.24 | 1.1 ± 1.3 | 0.222 |

BSA: Body surface area; STS: Society of thoracic surgeons
Results presented as mean ± standard deviation or number (%).
<sup>^</sup>Significant p-values presented in bold letters.

### Commissural alignment strategies

A CA technique was attempted in 215 patients (38.6%), including 113 patients with Acurate and 102 patients with Evolut series (**Central Figure**). The strategies for CA included specific orientation of the delivery system (at 6 o’clock for Acurate series and at 3 o’clock for Evolut series) in 135 patients (63.8%), and patient-tailored orientation as described by Redondo et al (8) in 80 cases (37.2%). The alignment was successful (none/mild misalignment) in 158 of them (73.5%). Angiographic evaluation of CMA degree demonstrated good correlation with its evaluation by CT (Kappa=0.9286 [0.8858-0.9715]; pondered Kappa=0.9719 [0.9544-0.9893]). No differences were found according to the device used (AN2: n=85 (75.2%); EP: n=73 (71.6%), p=0.545), but patient-tailored orientation demonstrated better rate of none/mild misalignment (n=72, 90%) than fix delivery system orientation following manufacturer instructions (n=86, 63.7%, p<0.001).

In those cases with no specific CA technique (342, 61.4%), none/mild CMA was achieved in 149 patients (43.6%), 140 (40.9%) presented moderate misalignment, and 53 (15.5%) severe misalignment. Therefore, the use of any technique (either with fix or patient-tailored orientation) demonstrated a statistically significant impact in the final rate of none/mild CMA (p<0.001), but no differences were found according to the device used (**Figure 1**). The analysis for coronary alignment could only be assessed for those patients with CT at follow up (n=226) and demonstrated commissural post-overlap with the right coronary artery ostium in 54 patients (23.9%) and with the left main in 38 patients (16.8%) of the global study population.

**Figure 1.**
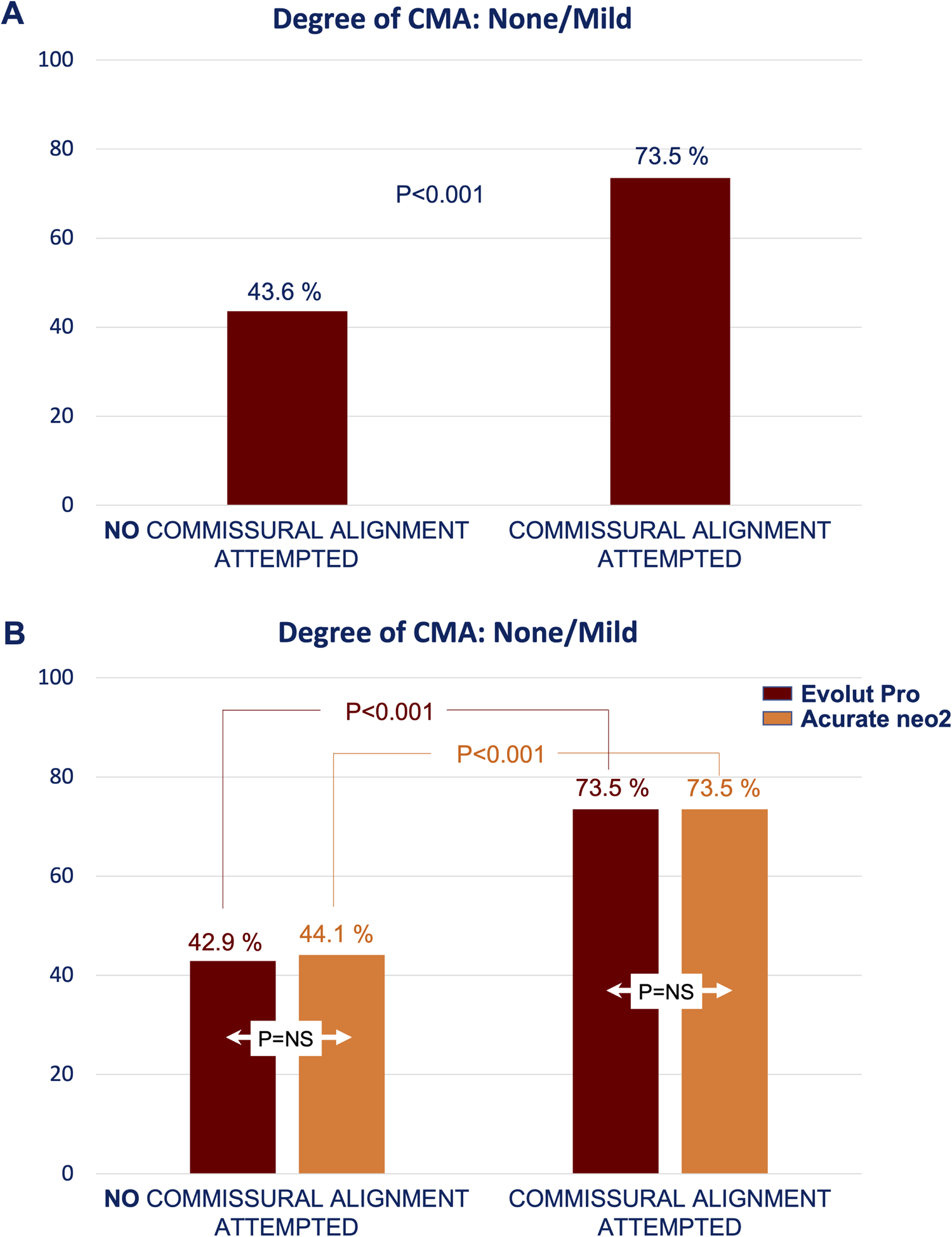
Rate of none or mild commissural misalignment according to the attempt or not to perform specific commissural alignment techniques and according to device. CMA: Commissural misalignment.

### Hemodynamic impact of commissural alignment

There were no significant differences at baseline regarding left ventricular function, aortic regurgitation degree, or the transvalvular mean gradients between patients who, after TAVR, presented CA and those who did not. However, peak gradient was higher at 30-day echocardiography in patients with moderate/severe CMA (21.8±8.8mmHg vs. 19.7±8.5mmHg, p=0.006). At 1-year follow up, differences persisted in peak gradient values (23.7±8.5mmHg vs. 22.3±8.8mmHg, p<0.001), and there was also a significantly greater progression of peak gradients in those with moderate or severe CMA that was confirmed for both, mean and peak transvalvular gradients’ progression after matching for baseline anatomical and hemodynamical confounders (**Table 3**). In the matched sample no differences were found between AN2 and EP devices (**Supp.Table 2**). Detailed report of mean and peak gradients according to the device used is summarized in **Figure 2**.

**Figure 2.**
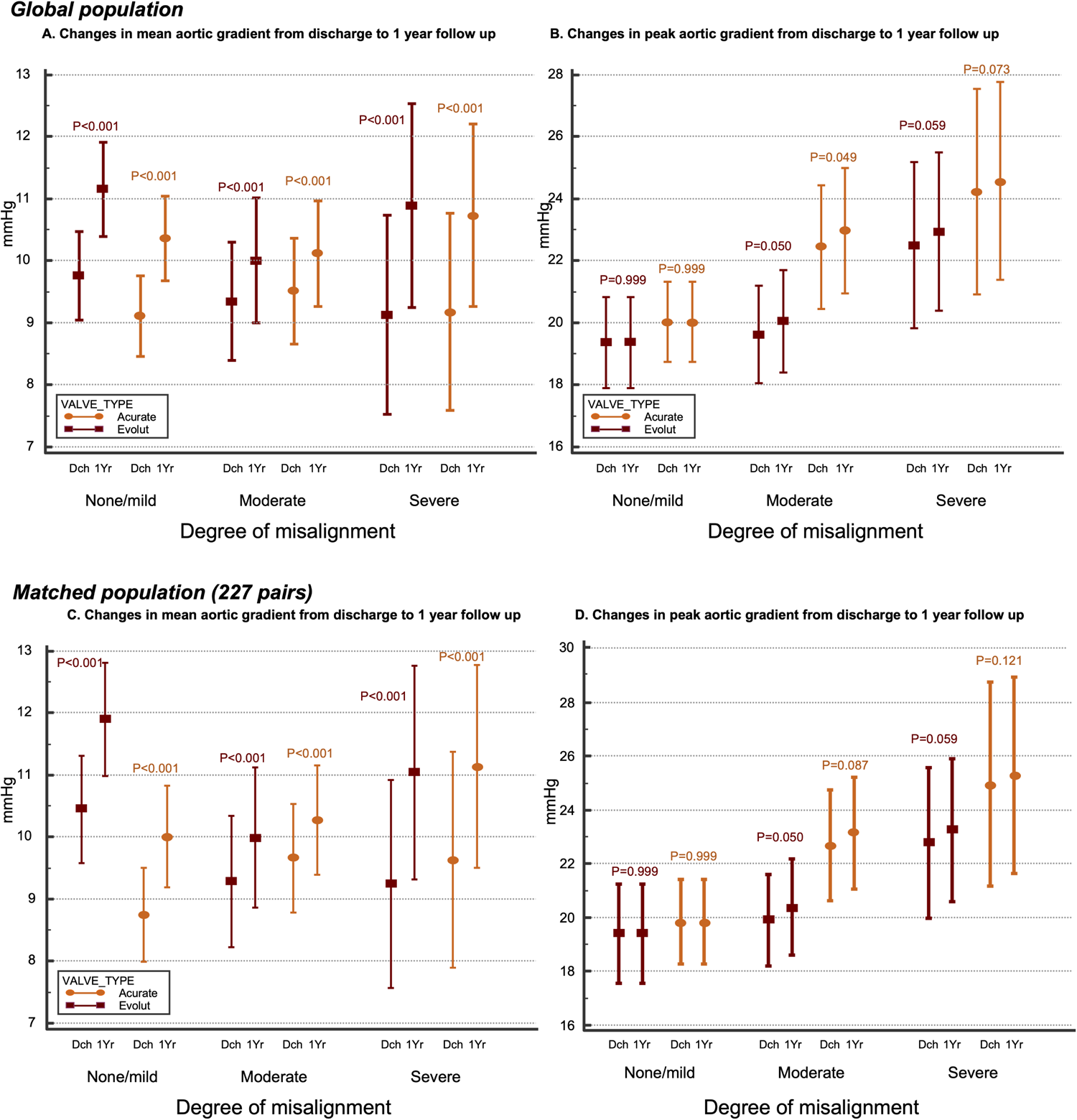
Progression of trans-prosthetic mean and peak gradients from discharge to 1-year follow-up according to the degree of commissural misalignment and to the device. Dch: Discharge. 1Yr: 1-year.

**Table 3.** Changes in trans-prosthetic aortic gradients from post-procedure to 1-year follow up according to the degree of misalignment. A: None/mild vs. moderate/severe misalignment. B: Non-severe vs. severe misalignment. C: None/mild vs. moderate/severe misalignment after matching for left ventricular ejection fraction, annulus diameter, eccentricity index, annulus perimeter, and type and size of the prosthesis.

| **A. Changes in trans-prosthetic aortic gradients from post-procedure to 1-year according to the degree of misalignment: None/mild vs. moderate/severe** |  |  |  |
| --- | --- | --- | --- |
|  | **None/mild CMA** | **Severe CMA** | **p-value<sup>^</sup>** |
| Mean gradient, mmHg (1-year FU) | 10.5 ± 4.6 | 10.2 ± 4.5 | 0.282 |
| *Δ mean gradient, mmHg* | 1.3 ± 1.2 | 1.2 ± 1.1 | 0.142 |
| Peak gradient, mmHg (1-year FU) | 19.7 ± 8.5 | 22.3 ± 8.7 | **0.001** |
| *Δ peak gradient, mmHg* | 0.1 ± 0.0 | 0.4 ± 0.2 | **<0.001** |
| **B. Changes in trans-prosthetic aortic gradients from post-procedure to 1-year according to the degree of misalignment: Non-severe vs. severe.** |  |  |  |
|  | **Non-severe CMA** | **Severe CMA** | **p-value<sup>^</sup>** |
| Mean gradient, mmHg (1-year FU) | 10.6 ± 4.6 | 10.8 ± 4.6 | 0.710 |
| *Δ mean gradient, mmHg* | 1.3 ± 1.2 | 1.7 ± 1.7 | 0.054 |
| Peak gradient, mmHg (1-year FU) | 19.7 ± 8.5 | 23.7 ± 8.5 | **<0.001** |
| *Δ peak gradient, mmHg* | 0.1 ± 0.0 | 0.4 ± 1.2 | **0.009** |
| **C. Changes in trans-prosthetic aortic gradients from post-procedure to 1-year according to the degree of misalignment: None/mild vs. moderate/severe\*.** |  |  |  |
| **\*Propensity score analysis matching for left ventricular ejection fraction, annulus diameter, eccentricity index, annulus perimeter, and type and size of the prosthesis.** |  |  |  |
|  | **None/mild CMA** | **Moderate/Severe CMA** | **p-value<sup>^</sup>** |
| Mean gradient, mmHg (1-year FU) | 10.8 ± 4.7 | 10.4 ± 4.5 | 0.352 |
| *Δ mean gradient, mmHg* | 0.9 ± 1.3 | 1.3 ± 1.3 | **0.001** |
| Peak gradient, mmHg (1-year FU) | 19.7 ± 8.9 | 22.5 ± 8.9 | **0.001** |
| *Δ peak gradient, mmHg* | 0.1 ± 0.1 | 0.4 ± 2.0 | **0.002** |

CMA: Commissural misalignment; FU: follow-up.
Results presented as mean ± standard deviation or number (%).
<sup>^</sup>Significant p-values presented in bold letters.

The proportion of patients with mean gradient ≥10mmHg at 1-year follow up, as a surrogate marker of structural valve deterioration, was similar irrespective of baseline degree of CMA (**Figure 3-A**) or the devices used (**Figure 3-B**).

**Figure 3.**
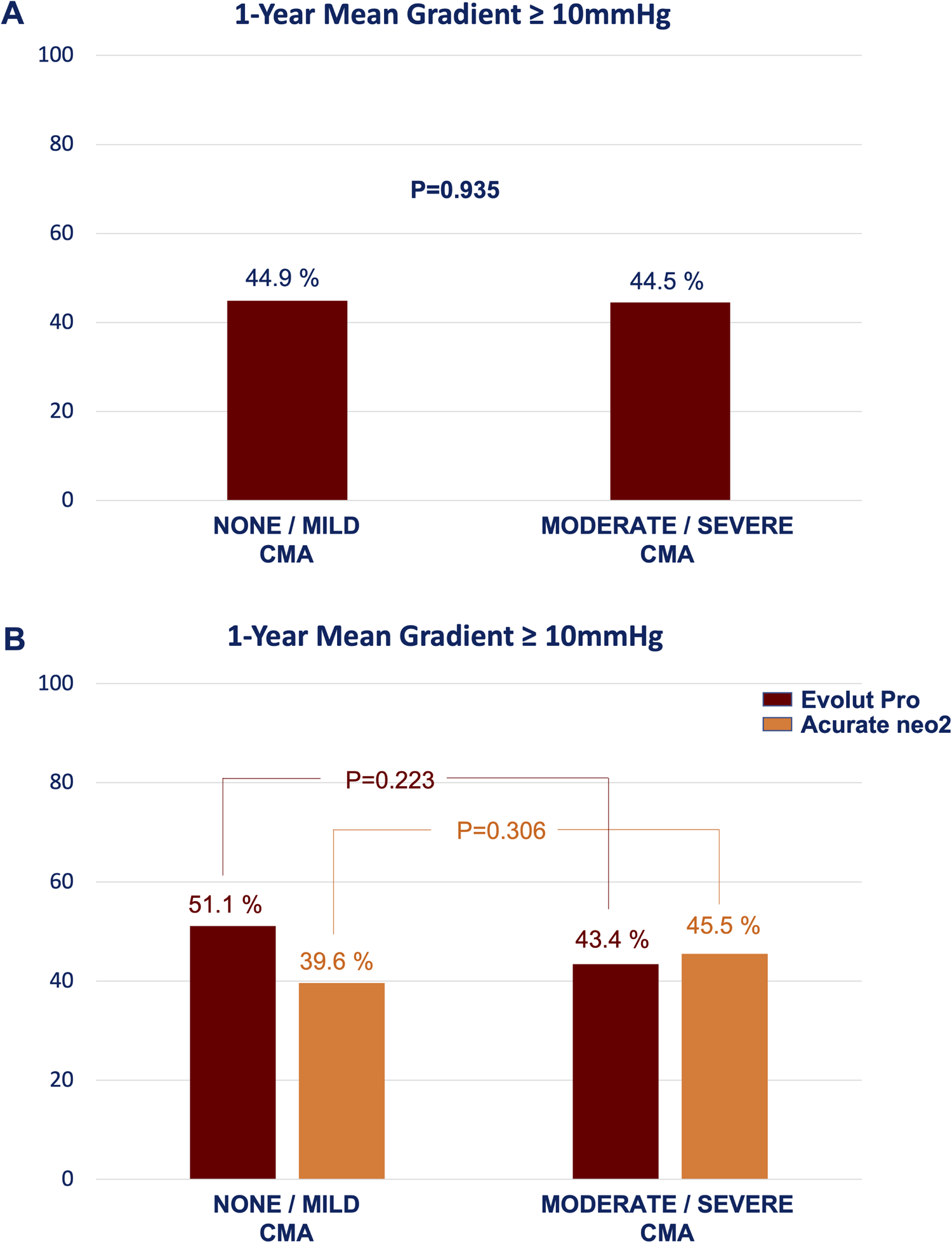
Rate of residual gradient ≥ 10 mmHg at 1-year follow up as indicator of structural valve deterioration according to the degree of commissural misalignment and to the device used. CMA: Commissural misalignment.

Up to one year of follow up, despite greater progression in the absolute number of patients with significant regurgitation amongst those with moderate/severe misalignment (12.4 vs. 8.5% in those with none/mild misalignment, p=0.200), there were no statistically significant differences in the aortic regurgitation severity following TAVR. However, the analysis of the mechanism of aortic regurgitation demonstrated that, the presence of moderate or severe intra-prosthetic leak was significantly higher in those without CA (1.2% vs. 0.4%, p=0.005) (**Figure 4**).

**Figure 4.**
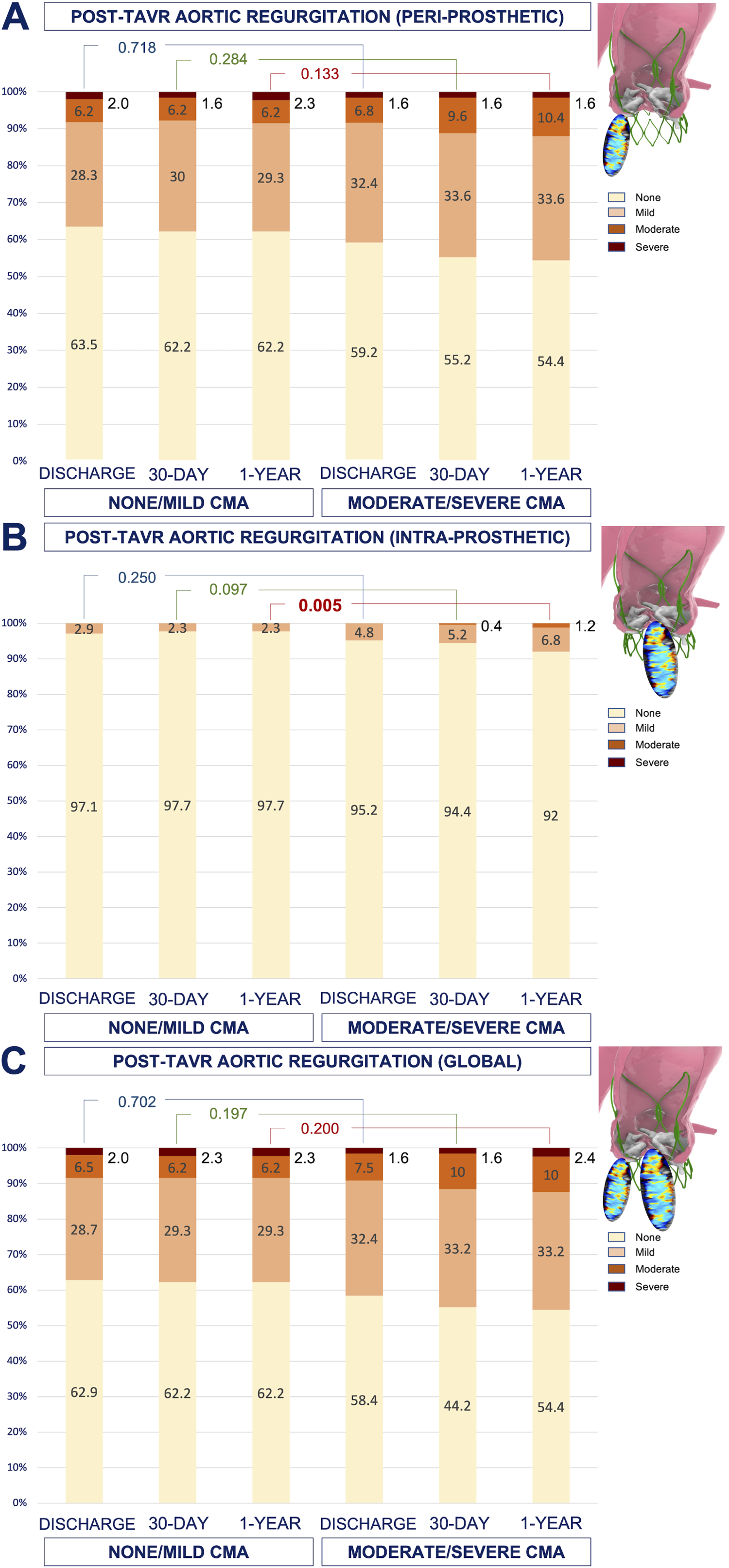
Rates of peri-prosthetic, intra-prosthetic, and global aortic regurgitation following TAVR during echocardiographic evaluation at discharge, 30-day, and 1-year according to the degree of commissural misalignment. CMA: Commissural misalignment. TAVR: Transcatheter aortic valve replacement.

## DISCUSSION

Commissural misalignment of TAVR can impair coronary re-access following supra-annular self-expandable devices (5) but might also have an impact on the hemodynamic behavior and, consequently, on the durability of the devices. In this regard, our research raised the following findings: 1/ CA techniques as a whole demonstrated efficacy in almost 3 out of 4 patients compared to only 43.6% when CA was not attempted; 2/ Patients with CMA presented a higher rate of aortic regurgitation at 1-year follow up with a significant increase of intra-prosthetic regurgitation; 3/ Finally, it was also observed that moderate/severe CMA was associated to higher transvalvular peak gradient and greater progression of both mean and peak gradients at 1-year after adjusting for main confounders although the proportion of patients with mean gradient ≥10mmHg remained similar irrespective of the degree of CMA. These findings represent a hint of a more rapid valve deterioration at 1-year of follow up although they do not meet the current definition of structural valve deterioration suggesting that longer-term follow up is crucial in this regard not only for balloon-expandable valves, but also for self-expandable devices despite the better acute gradients currently observed in clinical practice.

There is little bench test evidence regarding the hemodynamic effects of prosthetic CMA; recently, some investigation has suggested that the lack of CA is associated to increased shear stress of the leaflets (9). The RESOLVE study (1) after analyzing 324 patients also suggested that CMA was independently associated with an increase in transvalvular gradients at 30-day. No significant differences were found in terms of clinical outcomes, including death or stroke, or in the rate or progression of significant aortic regurgitation after TAVR. However, the balloon-expandable intra-annular prostheses Edwards SAPIEN 3 device (Edwards Lifesciences) presented a significant increase in gradients. The authors suggested that a potential greater recoil effect could occur in cases with CMA. But this mechanism is less likely with intra- or supra-annular self-expandable devices. Fuchs et al. (10) studied the possible impact of CA in a sample of patients that included supra-annular prosthesis, although together with surgical and intra-annular prosthesis. In this work, a greater central aortic regurgitation was also found in cases of moderate or severe misalignment, but the inhomogeneous sample precluded from specific conclusions regarding self-expandable TAVR. The trend to greater progression of aortic regurgitation in patients with moderate or severe CMA in our research could be explained by an impaired leaflet coaptation resulting from a more eccentric expansion of the device; it is known that annulus eccentricity conditions greater progression of regurgitation following self-expandable devices more often that with balloon-expandable prosthesis (11), which could be exacerbated in case of CMA. This aspect added to the more adverse fluid dynamics with greater shear stress on the edge of the leaflets (12,13) could lead to a less efficient bioprosthetic leaflet closure during early diastole and explain the increased rate of central leak in our sample. Conversely, this eccentricity of the annulus is unlikely to be a contributing factor to the increased gradients progression given that it persisted after the matched comparison that included annular dimensions and eccentricity, but it cannot be excluded its relationship with the altered shear stress as has been previously suggested in computational models (12). Recent research by Jung et al (13) suggested that hypo-attenuated leaflet thickening (HALT) was more often detected among subjects with greater deviation of the commissures and, indeed, severe CMA resulted an independent predictor of HALT after TAVR (OR=2.2, p=0.018). Computational models have previously failed to demonstrate an increased platelet activation in this context (12) but blood flow stagnation has already been describe for balloon-expandable TAVR devices (1). Conversely, the presence of HALT was detected in only 5 patients (3 with mild and 2 with moderate CMA) in the group of patients from our sample with CT at follow up. To remark, most of the studies were performed at 1-year of follow up in our sample as opposed to the former investigation where some of the patients were evaluated at 3-month follow up. In addition, no differences were found by the authors in the proportion of cases with HALT treated with balloon- or with self-expandable devices. It is plausible that this mechanism, through endothelial dysfunction and alteration in blood flow patterns, occurred also in our population and could explains the detection of an increased progression of gradients; still the clinical relevance remains unknown since no differences were detected in mortality, stroke or other major clinical outcomes. Finally, imaging studies including magnetic resonance, have suggested from the early stages of TAVR technology the risk of blood stasis in the neo-sinuses; the commissural posts centered in the mid part of the sinuses of Valsalva might occupy and divide this part of the aortic root increasing the difficulty for high-speed continues blood flow, particularly with supra-annular devices, and therefore leading to thrombus formation (14,15).

The main limitations of this work include the ambispective design and the variability of techniques used to explore the degree of CA, although angiography and CT correlated well. Despite the recent consensus by the ALIGN-TAVR consortium, still certain variability in the definition of the degrees of CA exists and, in fact, the current threshold and the 4 different categories are an empirical classification that might correlate poorly with actual clinical events in the follow up. Data on clinical and hemodynamic outcomes were only available at 1-year follow up, which might be short to determine eventual greater impact of CMA.

## CONCLUSIONS

CA strategies can be effectively used for self-expandable TAVR devices presenting better results compared with traditional uncontrolled orientation of TAVR delivery systems. Still, in more than 25% of the cases CA techniques failed and moderate or severe CMA was associated to a greater progression of central aortic regurgitation and transvalvular gradients at 1-year follow up. Despite the lack of clinical impact and the unmet criteria for structural valve deterioration, these findings after relatively short follow up represent a hint on the true impact of CMA in valve durability and warrant longer term follow up to clarify this aspect.

## Data Availability

Data are available for other investigators for further research under request.

## ABBREVIATIONS

AN2: Acurate neo2.
CA: Commissural alignment.
CMA: Commissural misalignment.
CT: Computed tomography.
EP: Evolut Pro
TAVR: Transcahteter aortic valve replacement.

---

**SUPPLEMENTAL FIGURE 1.**
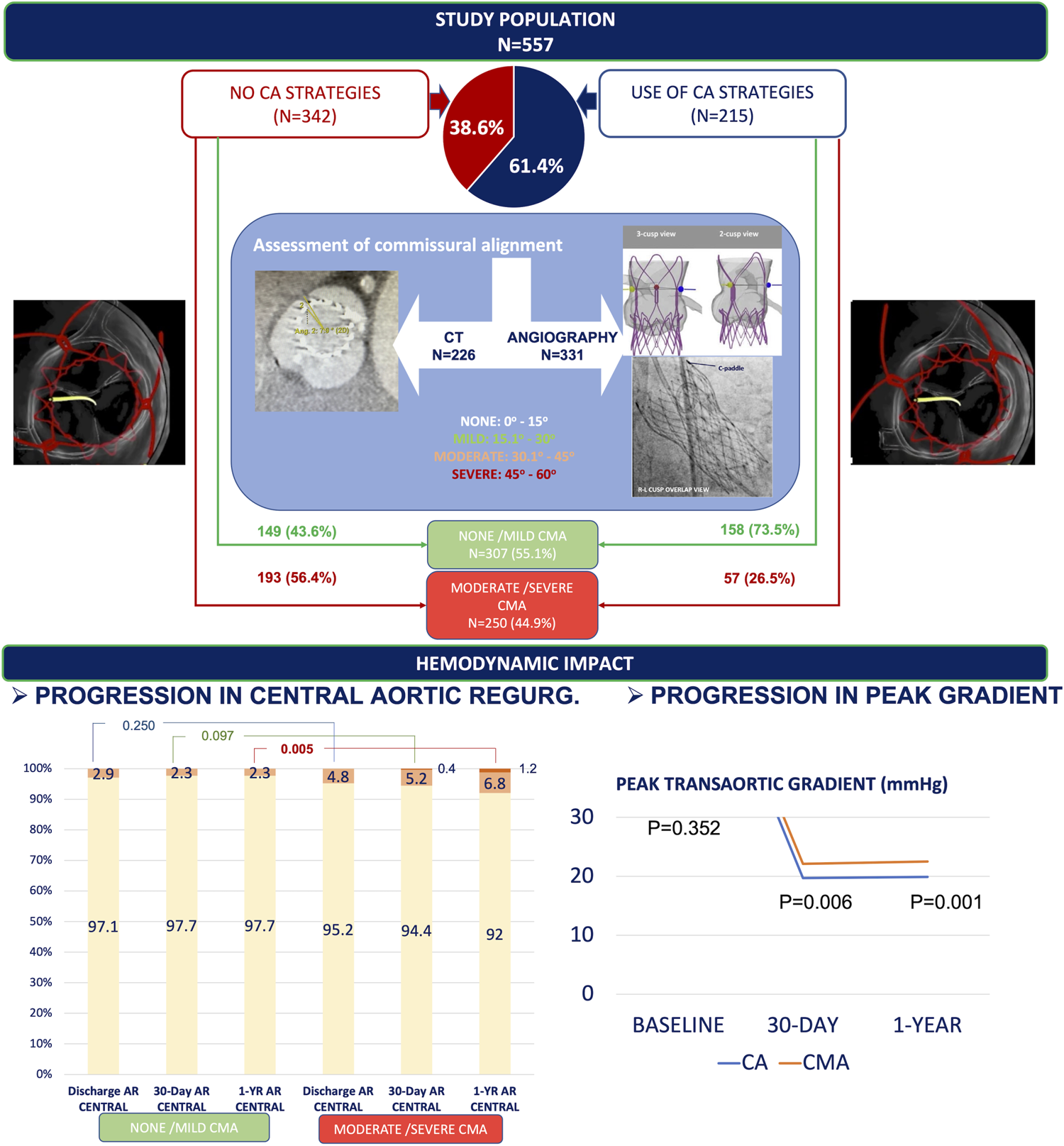
Degree of adjustment of variables included in the matched comparison.

